# How is sleep-disordered breathing linked with biomarkers of Alzheimer’s disease?

**DOI:** 10.1101/2023.08.16.23294054

**Authors:** Mohammad Akradi, Tara Farzane-Daghigh, Amir Ebneabbasi, Hanwen Bi, Alexander Drzezga, Bryce A. Mander, Simon B. Eickhoff, Masoud Tahmasian, the Alzheimer’s Disease Neuroimaging Initiative

## Abstract

Sleep-disordered breathing (SDB) is prevalent in Alzheimer’s disease (AD). We assessed whether and how SDB affects neuroimaging biomarkers of AD, including amyloid-beta plaque burden (Aβ), regional uptake of fluorodeoxyglucose using positron emission tomography (rFDG-PET), grey matter volume (GMV), as well as cognitive scores and cerebrospinal fluid (CSF) biomarkers.

**Methods:** We selected 757 subjects from the Alzheimer’s Disease Neuroimaging Initiative (ADNI) database based on cognitive status (AD, mild cognitive impairment (MCI), cognitively unimpaired (CU)), and SDB condition (with/without SDB). To ensure the reliability of our findings and considering imbalanced sample size across groups, we used a stratified subsampling approach generating 10,000 subsamples (n=10/group). We then selected 512 subsamples with matched covariates. The effect size of the cognitive status-SDB interaction was computed for each biomarker and cognitive score. For reference, we computed 1000 null models by shuffling group labels randomly. The average value of effect sizes for each biomarker in each region was estimated through bootstrapping with 10,000 iterations for both the main and null models and compared with the null model’s distribution. Linear regression models were next implemented to identify associations between the effect size on Aβ, rFDG, and GMV with the effect size on cognitive scores and CSF biomarkers across all subsamples.

**Results:** The cognitive status-SDB interaction had a medium-sized effect on Aβ, rFDG and GMV biomarkers in several brain areas. The effect sizes of the mentioned interactions on Aβ plaque burden in the right precuneus, left middle temporal gyrus, and left occipital fusiform gyrus were associated with the effect sizes of the interactions on cognitive scores. Further, the interaction effect sizes on CSF Aβ42 were related to the interaction effect sizes on Aβ in the right precuneus and posterior cingulate cortex, as well as rFDG in the left precuneus cortex and GMV in bilateral angular gyrus and right occipital fusiform gyrus. Effect sizes on CSF p-tau were also correlated with the effect sizes on Aβ in the left lateral occipital cortex and GMV in the left middle temporal gyrus.

**Conclusion:** We observed that SDB interacts with neuroimaging and CSF biomarkers of AD. Specifically, SDB has a robust association with markers of Aβ pathology in PET and CSF relative to rFDG and GMV in the AD group. The cognitive status-SDB interaction on Aβ is associated with cognitive decline. This study further supports the hypothesis that SDB may precipitate AD pathology.

## Introduction

Alzheimer’s Disease (AD) constitutes 60–70% of all cases of dementia, making it the most common form of neurodegenerative disease worldwide^1^. Recently, it has been estimated that by 2050, the prevalence of AD will be three-fold and, thus, affect about 50 million people worldwide^2,3^. AD is one of the most lethal, costly, and burdensome diseases in the 21st century^4^. Cognitive and behavioral functions, including episodic memory, language, attention, reasoning, and judgment, are progressively impaired in AD^5^. Identification of risk factors for AD is critical, e.g., it has been shown that healthy lifestyle habits (e.g., regular physical activity) and management of cardiovascular risk factors may reduce the risk for dementia^6^. The modifiable risk factors for AD include diabetes, obesity, hypertension, vascular disease^7^, and sleep disorders such as insomnia^8^ and Sleep-Disordered Breathing (SDB)^9^. SDB is an umbrella term characterized by abnormal respiration during sleep that ranges from heavy and loud snoring to frequent episodes of partial or complete airway obstruction for at least 10 seconds; so-called Obstructive Sleep Apnea (OSA), OSA is the most common form of SDB, which leads to intermittent hypoxemia, arousal, and sleep fragmentation^10^. SDB is widely prevalent and affects 52.6% of men and 26.3% of women over 60 years old^11^, although it is often underdiagnosed, especially in women^12^. and. Lower quality of life, a higher risk of road traffic accidents, and various health conditions, including cardiovascular problems, metabolic disorders, and cognitive impairments, are often seen in patients with SDB^13^. Thus, assessing the pathophysiology of SDB and its consequences in the general population is critical.

Recent studies suggest a complex but strong relationship between SDB and AD^14–17^. A meta-analysis found that patients with AD have a 5-fold increased risk of presenting with SDB compared to cognitively normal individuals^9^. SDB advances cognitive impairment in individuals with Mild Cognitive Impairment (MCI) or AD, and such cognitive decline happens earlier in the affected patients^18^. Hence, patients with SDB symptoms are more likely to develop MCI or AD dementia in the future ^16,18,19^. Transgenic AD mouse models found that the induction of chronic intermittent hypoxia, which is a characteristic of SDB in human, increased brain Amyloid-beta (Aβ)^20^ plaque burden and tau phosphorylation^21^. A longitudinal study found that baseline SDB severity was associated with increased cortical amyloid burden in cognitively normal older adults^22^. Another study observed higher cortical Aβ accumulation in middle-aged patients with severe SDB^23^. Further, it was revealed that in healthy and MCI groups, Cerebrospinal Fluid (CSF) Aβ decreased, and CSF phosphorylated tau (p-tau) increased, particularly in subjects with SDB^24^. Thus, SDB may interact with AD pathophysiology in preclinical stages to increase the risk of developing AD and finally exacerbate the development of AD-related cognitive impairments.

Developing biomarkers, including amyloid-beta Positron Emission Tomography (Aβ-PET), 18F-Fluorodeoxyglucose PET (FDG-PET), and structural Magnetic Resonance Imaging (sMRI)^25^, as well as Aβ and tau assessment in CSF, advanced our understanding of pathophysiological mechanisms, early detection, and monitoring of the progress and treatment of AD^26^. The recently introduced framework (i.e., Amyloid-Tau-Neurodegeneration AT(N)) classified biomarkers into Aβ deposition, pathologic tau, and neurodegeneration^27^. Focusing on these biomarkers may help to understand the underlying neurobiological mechanisms of the association of SDB with AD progression. Andre and colleagues observed that SDB was associated with medial temporal lobe atrophy in cognitively normal older adults, mainly in amyloid-positive individuals^28^. However, another study did not observe any association between SDB and structural brain alterations and brain aging in AD patients^29^. Higher Grey Matter Volume (GMV), Aβ, brain perfusion, and glucose metabolism, mainly in the posterior cingulate cortex (PCC) and precuneus, have been reported in SDB-positive subjects. However, such brain alterations were not associated with cognitive decline^30^. A recent systematic review summarized the inconsistent GMV and FDG-PET findings on the association between SDB and AD, highlighting the need for a comprehensive multimodal approach focusing on AD biomarkers in patients with SDB^16^.

In the present cross-sectional study, we assess the association between SDB presence with Aβ PET imaging (as a proxy of Aβ plaque burden in the brain), regional uptake of fluorodeoxyglucose metabolism (rFDG) indicating cerebral glucose metabolism (as a surrogate of neural activity), and GMV (as an indicator of brain structure) of cortical and subcortical brain areas along the trajectory of AD (i.e., cognitively unimpaired (CU), patients with MCI and AD). In particular, our questions were the following: (a) does the interaction between cognitive status (CU, MCI, AD) and SDB condition (SDB+, SDB-) affect Aβ, rFDG, and GMV; (b) are the candidate brain region(s) affected by SDB linked with cognitive impairment. To address these knowledge gaps, multimodal neuroimaging and CSF data from the Alzheimer’s Disease Neuroimaging Initiative (ADNI) were used. In the complementary analyses, the probable relationships between neuroimaging and CSF biomarkers of AD, including Aβ42 and p-tau were assessed. Over the last few years, it has become apparent that neuroimaging research faces a replication crisis that has prompted the collection of big data samples and replication studies to reduce false-positive rates and increase the generalizability of findings^31–33^. Thus, to increase the robustness of results and considering our imbalanced sample size across our groups, we have divided our individuals into well-matched subsamples (n=10) in terms of covariates (e.g., age, gender, APOE ε4, BMI), and we investigated how interactions between cognitive status and SDB conditions are associated with AD biomarkers.

## Materials and methods

### Participants

Data were obtained from the ADNI database (adni.loni.usc.edu)^34^. We selected participants with the three-mentioned imaging modalities, including 18F-AV45 (florbetapir) amyloid-beta PET, FDG-PET, and sMRI, and Mini-Mental State Examination (MMSE) as a measure of subjects’ cognitive performance, as well as CSF measures of Aβ42 and p-tau. Subjects with a history of stroke, Parkinson’s disease, or brain tumors were excluded. PET and sMRI scans and MMSE assessment data were collected within two months of each other. Diagnoses of the subject’s cognitive status were based on the ADNI reports. From 757 included subjects, the 102 “SDB+” subjects were defined with the self-reported “sleep apnea”, and/or history of “obstructive sleep apnea” “OSA” or “sleep-disordered breathing” in the ADNI dataset. Otherwise, participants without any self-reported SDB symptoms are considered as “SDB-”, which included 655 participants. The SDB+ individuals who had received any treatment like Continuous Positive Airway Pressure (CPAP) or bi-level positive airway pressure (BiPAP or BPAP) were excluded, as it has been shown that CPAP treatment may minimize the effects of SDB on biomarkers of AD and cognitive decline^18^, as well as CSF Aβ42 and p-tau^35^. Furthermore, MMSE, CSF Aβ42 and p-tau data were obtained from the subject’s medical history. After obtaining and labeling data, we assembled six distinct groups according to their cognitive status (CU, MCI, AD) and SDB condition (SDB+ or SDB-; Table 1).

**Table 1.** Characteristics of the study participants. (*) cerebrospinal fluid. SDB: sleep-disordered breathing.

|  | **SDB-** | **SDB+** |
| --- | --- | --- |
| **Alzheimer's Disease (AD)** | **N: 114** | **N: 16** |
| Age (mean [SD]) | 75.1 (8.1) | 70.2 (6.5) |
| Age range | [55.7, 90.4] | [60.8, 82.2] |
| Sex, female (%) | 48 (42%) | 6 (38%) |
| Body-mass index (mean [SD]) | 25.5 (4.31) | 27.3 (4.67) |
| CSF* Aβ 42 (mean [SD]) | 823.38 (476.7) | 610.25 (215.6) |
| CSF* p-tau (mean [SD]) | 37.13 (15.8) | 35.59 (14.23) |
| Education years (mean [SD]) | 15.77 (2.64) | 15.75 (2.52) |
| Aβ-positive (%) | 101 (88.6%) | 14 (87.5%) |
| **$\geq 1$ APOEε4 allele presence (%)** |  |  |
| False | 37 (33%) | 3 (19%) |
| True | 76 (67%) | 13 (81%) |
| missing | 1 | 0 |
| MMSE (mean [SD]) | 23.04 (2.11) | 22.94 (1.98) |
| **Mild Cognitive Impairment (MCI)** | **N: 276** | **N: 58** |
| Age (mean [SD]) | 72.23 (7.58) | 71.24 (7.58) |
| Age range | [55.1, 91.5] | [55.1, 88.5] |
| Sex, female (%) | 130 (47%) | 18 (31%) |
| Body-mass index (mean [SD]) | 26.67 (5.36) | 29.18 (6.22) |
| CSF* Aβ42 (mean [SD]) | 1101.153 (565.39) | 1241.29 (572.41) |
| CSF* p-tau (mean [SD]) | 28.23 (15.09) | 24.1 (12.82) |
| Education years (mean [SD]) | 16.3 (2.6) | 16.22 (2.79) |
| Aβ-positive (%) | 258 (93.5%) | 52 (89.7%) |
| **$\geq 1$ APOEε4 allele (%)** |  |  |
| False | 125 (46%) | 35 (64%) |
| True | 148 (54%) | 20 (36%) |
| missing | 3 | 3 |
| MMSE (mean [SD]) | 27.69 (2.48) | 27.93 (1.57) |
| **Cognitively Unimpaired (CU)** | **N: 265** | **N: 28** |
| Age (mean [SD]) | 74.36 (6.41) | 72.87 (5.51) |
| Age range | [56.3, 93.8] | [65.2, 85.7] |
| Sex, female (%) | 157 (59%) | 10 (36%) |
| Body-mass index (mean [SD]) | 27.33 (5.22) | 29.19 (5.08) |
| CSF* Aβ42 (mean [SD]) | 1438.58 (639.23) | 1755.21 (661.32) |
| CSF* p-tau (mean [SD]) | 22.74 (10.04) | 21.58 (9.73) |
| Education years (mean [SD]) | 16.52 (2.48) | 16.46 (2.95) |
| Aβ-positive (%) | 237 (89.4%) | 25 (89.3%) |
| **$\geq 1$ APOEε4 allele (%)** |  |  |
| False | 188 (72%) | 22 (88%) |
| True | 75 (28%) | 3 (12%) |
| missing | 2 | 3 |
| MMSE (mean [SD]) | 29.01 (1.23) | 29.07 (1.02) |

### Imaging acquisition and preprocessing

For PET images, a detailed explanation of the image acquisition is available on the ADNI website (http://adni.loni.usc.edu/pet-analysis-method/pet-analysis). All PET and structural images underwent a primary preprocessing protocol by ADNI. Briefly, PET volumes were averaged, spatially aligned, and interpolated to the standard voxel size (1.5×1.5×1.5 mm). Furthermore, we preprocessed all images using FMRIB software library (FSL)^36^. To match inter-modality intra-subject data for each subject, both PET and sMRI (T1) images were co-registered into the MNI152 space nonlinearly using the FNIRT pipeline in FSL^37^. Then, T1 images were normalized to the MNI152 template with affine transformations, and PET images were registered to T1 images with rigid body transformations. Next, PET images were registered into MNI152 space using the obtained transformation matrixes. Finally, preprocessed PET images were scaled by normalization of voxel-wise Aβ and rFDG uptake values to the cerebellar vermis Aβ and rFDG uptake, respectively, as applied previously^38–40^. For 100 cortical and 16 subcortical regions, rFDG and Aβ values were extracted using an obtained map according to the Schaefer cortical^41^ and Melbourne subcortical^42^ parcellations.

Participants underwent a standardized protocol for high-resolution T1 MRI brain scans^43^. T1 imaging acquisition parameters were: TR = 2,400ms, minimum full TE, TI = 1,000ms, 24 cm field of view, acquisition matrix of 192 × 192 × 166 and with 1.25×1.25×1.2 mm3 slice size (for more detail, see https://adni.loni.usc.edu/methods/mri-tool/mri-analysis). To perform voxel-based morphometry (VBM), we used an FSL-VBM protocol^44^. As the first step, the brain extraction tool and grey matter segmentation were applied to the structural images before being registered to the MNI 152 standard space using non-linear registration^37^. To create a study-specific grey matter template, the resulting images were averaged and flipped along the x-axis. Secondly, all native T1 images were non-linearly registered to the study-specific template and modulated to correct for local expansion (or contraction) due to the non-linear component of the spatial transformation^41^. Finally, as described above, GMV scores of the 100 cortical and 16 subcortical regions were also extracted.

### Stratified subsampling

To achieve robust and generalizable findings and in order to minimize the impact of covariates such as age, sex, APOE ε4^45^, and BMI^46^, out of 10,000 random stratified subsampling, 512 matched subsamples were selected such that there was no significant difference in age, gender, BMI, and presence of APOE ε4 between cognitive status and SDB condition levels. The number of subjects per subgroup was set as 10 for the reshuffling of subsamples due to the fact that the AD+SDB subgroup had only 16 subjects in total. The non-significant differences for covariates were checked by t-test for continuous variables (age and BMI; p > 0.05) and chi-square test for categorical variables (gender and presence of APOE ε4; p > 0.05). All the following analyses were applied to these 512 subsamples.

### Analysis of covariance

To evaluate the interactive effects of cognitive status and SDB condition on neuroimaging biomarkers of AD, a two-way ANCOVA was performed with cognitive status and SDB condition as main factors, while age, gender, APOE ε4 presence, and BMI were considered as covariates of no-interest. Then, the partial Eta-squared method was adopted to calculate the cognitive status-SDB interaction effect size on Aβ, rFDG, and GMV^47^. Next, a two-way ANCOVA was performed with MMSE-score as the dependent variable, and partial Eta-squared for cognitive status-SDB interaction was estimated. Two complementary ANCOVAs were performed to estimate partial Eta-squared of cognitive status-SDB interaction on CSF Aβ42 and CSF p-tau. This procedure was performed for Aβ, rFDG, and GMV across all subsamples, leading to an average value of 512 estimated effect sizes (using bootstrapping with 10,000 iterations) for each biomarker in each region to select medium effect-size measurements. All statistical analyses were computed in R version 4.2.0. The 95% confidence intervals and the mean values of calculated effect sizes for all subsamples were estimated for each parcel and biomarker.

### Null model estimation

For reference, we designed a null model, which was randomly estimated by shuffling cognitive status and SDB condition labels through participants. Then, the ANCOVA tests were replicated to obtain effect sizes of cognitive status-SDB interaction on each parcel and each biomarker in all subsamples. The bootstrapped average of effect sizes for each parcel and biomarker was estimated to be the same as the main model for all 512 subsamples. This procedure was repeated 1000 times, and the distribution of null effect size mean was estimated for each parcel and each biomarker. Afterward, the null distribution was used in order to test estimated effect sizes from the previous stage.

### Bootstrapping

As part of our analysis, we used a bootstrapping technique to estimate the average value of each biomarker in each region using 512 subsamples. In this approach, we sampled the data randomly with replacement, calculated each iteration’s mean, and aggregated the mean values. The bootstrapped means were calculated 10,000 times, yielding a distribution of averages and lower and upper confidence intervals that correspond to the 95% confidence intervals. Finally, for the main model, we estimated the average value of bootstrapped means. However, in the null model, after 1000 iterations, a distribution of bootstrapped mean values was obtained.

### Group comparisons

In order to explore the nature of the cognitive status-SDB interaction, we conducted group comparisons focusing on regions with medium effect sizes. Regions were defined by an effect size threshold greater than 0.04 and exceeding the null distribution maximum value. We applied a t-test to determine if there were significant differences in neuroimaging biomarkers between the SDB+ and SDB-groups among the subjects based on the average values of the biomarkers within these regions. This analysis further complemented our previous analyses, providing additional insights into the interaction between cognitive status and SDB condition.

### Linear regression

To reveal a potential association between neuroimaging and cognitive variables^48^, MMSE effect sizes were considered as a response, and all medium effect sizes of neuroimaging biomarkers were considered as explanatory variables. Explanatory variables with p < 0.05 were then significantly associated with the response variable. This linear regression approach aimed to estimate how much of the variance in the effect sizes on MMSE was explained by the variance in the effect sizes on neuroimaging biomarkers. Likewise, two more linear regressions were applied, once with CSF Aβ42 and once with CSF p-tau as dependent (response) variables, to investigate whether there is any link between effect sizes of neuroimaging biomarkers with CSF Aβ42 and CSF p-tau.

### Data availability

The dataset utilized in this research is owned by the Alzheimer’s Disease Neuroimaging Initiative (ADNI), a neuroscience collaboration between universities and research institutions. The ADNI database (<u>adni.loni.usc.edu</u>) offers access to the data after obtaining approval for a data request application. Additional details on accessing ADNI data can be found at https://adni.loni.usc.edu/datasamples/accessdata. A list of selected data for this study with their RID, sex, age, weight, cognitive status, SDB condition, and MMSE score at the time of neuroimaging data acquisition is provided in Supplementary Table 4.

## Results

Our subsampling technique, which involves selecting 512 matched subsamples out of 10,000, ensures that our results are not biased by sampling. This subsampling method, together with the use of Bootstrap to estimate the mean biomarker value, improves the stability and generalizability of our results. In addition, we conducted a null model estimation to establish a reference for comparison and validation of observed effect sizes. By comparing our effect sizes with the null effect size distribution, we can confidently assess the significance of the results and distinguish them from random variations.

By emphasizing these reliable measures in the results section, we highlight new aspects of our study, distinguishing them from previous analyses. The subsampling, bootstrapping, and null model estimation approaches used in our analysis provides a solid foundation for the interpretation and implications of our findings. For each neuroimaging biomarker in each region, the bootstrapped average value of cognitive status-SDB interaction effect sizes and the null model distribution are illustrated in Supplementary Fig. 1. To report regions’ names in this section, we used the Anatomy Toolbox to map the parcels from the Schaefer atlas to the standard neuroanatomical labels^49^.

### The association between cognitive status-SDB and Aβ plaque burden

The cognitive status-SDB interaction had a medium effect size on Aβ plaque burden in the left middle temporal gyrus, left angular gyrus, left inferior frontal gyrus, left cingulate gyrus, right frontal pole, bilateral superior frontal gyrus, right precuneus cortices, left occipital fusiform gyrus, and left lateral occipital cortex. Among subcortical regions, Aβ within both the posterior thalamus and the left nucleus accumbens was associated with a cognitive status-SDB interaction (Fig. 1A, Supplementary Fig. 1). A main effect of SDB condition also had a medium effect size on Aβ within the left frontal pole (Supplementary Fig. 2). In the AD group, in the regions that showed a medium-effect size of the cognitive status-SDB interaction, Aβ plaque burden was higher in the SDB+ than the SDB-subjects. However, the Aβ burden was lower in the SDB+ group than the SDB-group in both CU and MCI groups (Fig. 2A).

**Fig. 1.**
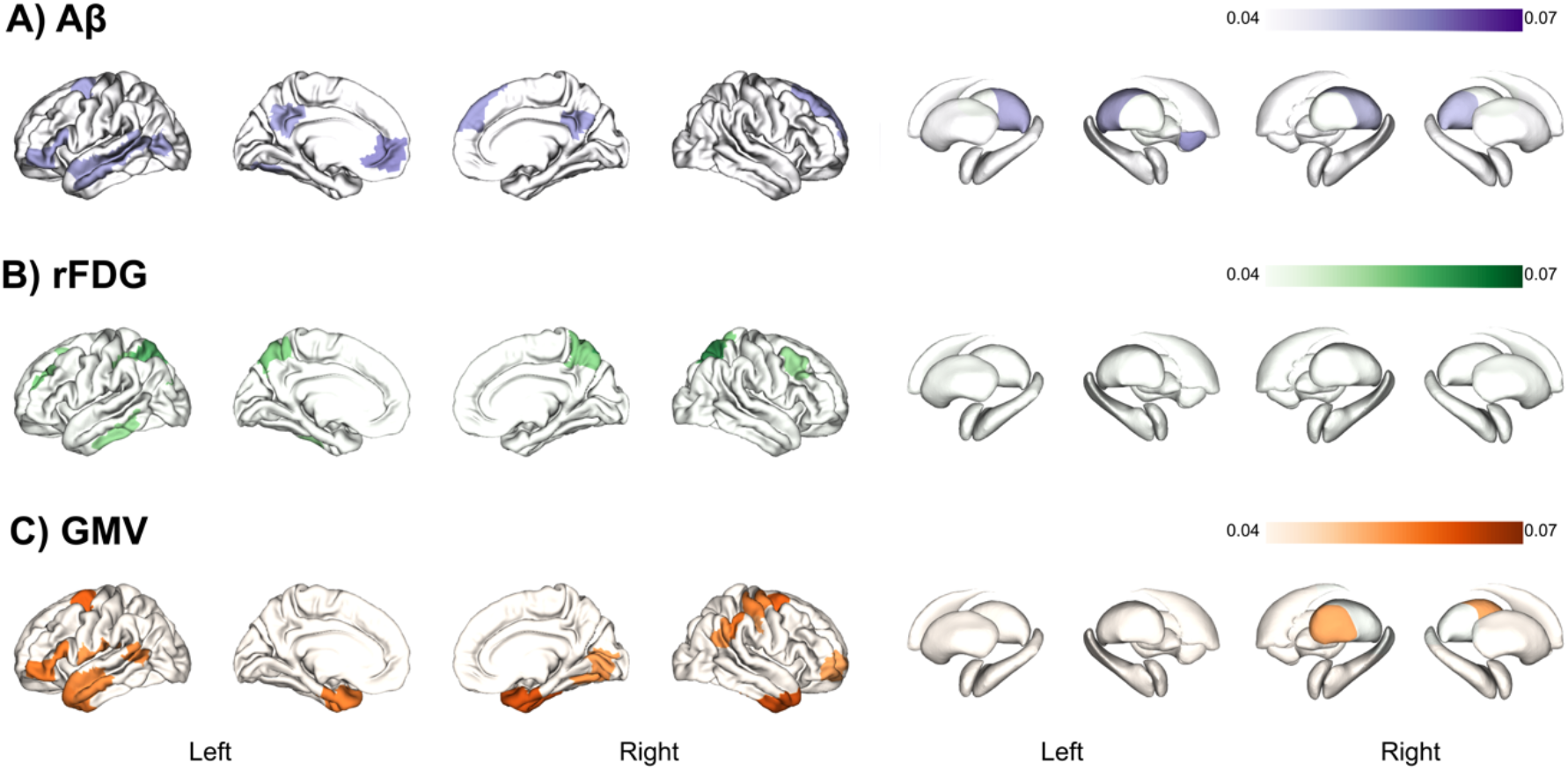
Cognitive status-SDB interaction medium effect sizes on AD biomarkers. The brain regions that have medium effect sizes of the interaction between cognitive status (Alzheimer’s disease, mild cognitive impairment and cognitively unimpaired) and SDB condition (with and without sleep-disordered breathing) for different biomarkers: A) amyloid-plaque burden (Aβ), B) regional fluorodeoxyglucose (rFDG), C) grey matter volume (GMV).

**Fig. 2.**
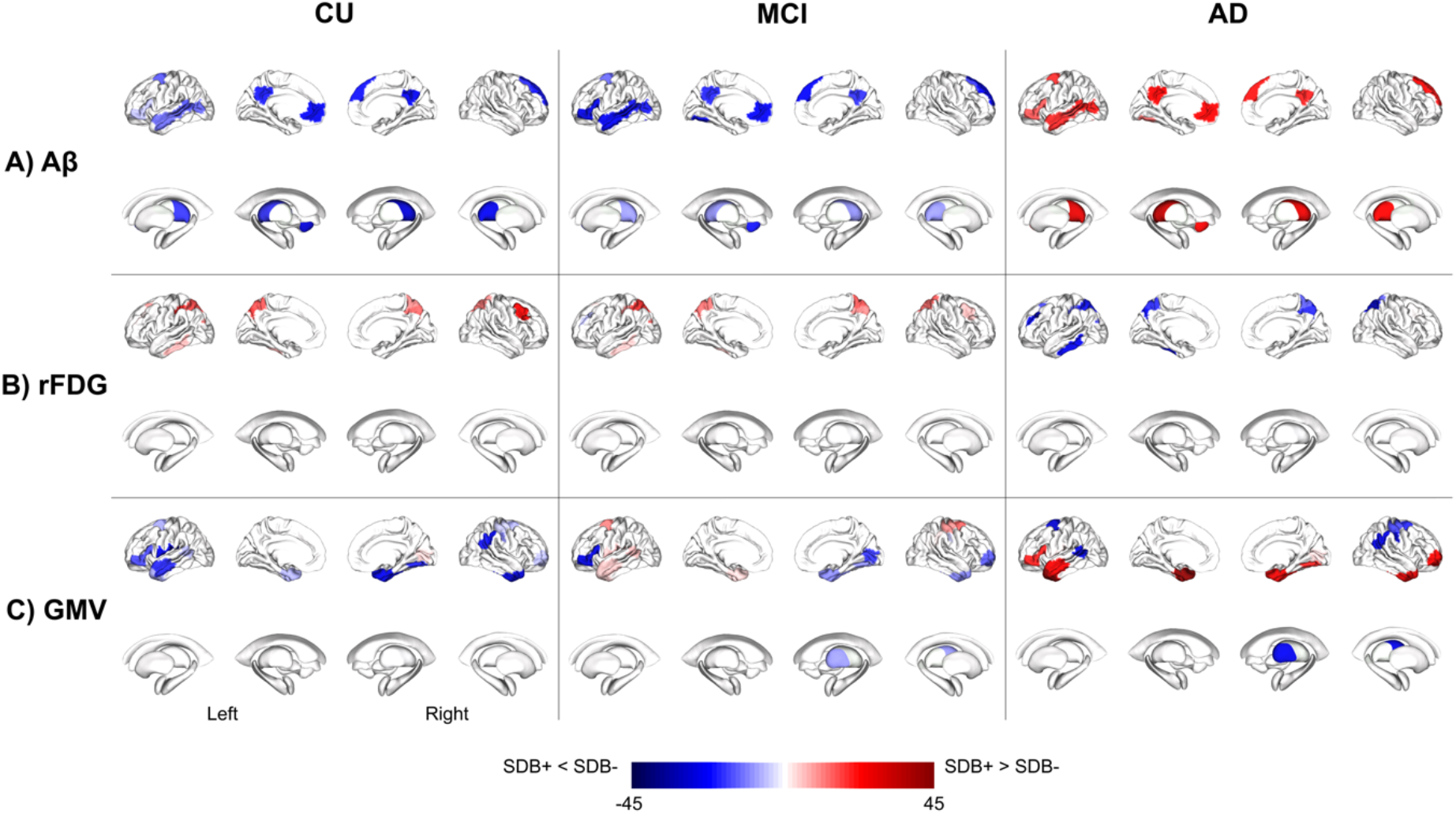
AD biomarkers alteration in different groups. Differences in each biomarker value in different states of SDB condition (with and without SDB) through different disease groups (Alzheimer’s disease, mild cognitive impairment, and cognitively unimpaired).

### The association between cognitive status-SDB and glucose metabolism

The cognitive status-SDB interaction was linked with regional glucose metabolism in the left postcentral gyrus, bilateral superior parietal lobule, bilateral lateral occipital cortex, left frontal pole, left inferior temporal gyrus, right middle frontal gyrus, bilateral precuneus cortex, and left superior frontal gyrus (Fig. 1B, Supplementary Fig. 1). A main effect of SDB condition alone, had a medium effect size on rFDG in the left precentral gyrus (Supplementary Fig. 2). In addition, in patients with AD, rFDG was lower in SDB+ compared to SDB-group but higher in the CU or MCI groups in all regions where rFDG was affected by the cognitive status-SDB interaction (Fig. 2B).

### The association between cognitive status-SDB and grey matter volume

A medium effect size was detected for the cognitive status-SDB interaction on GMV measures in the left central opercular cortex, right precentral gyrus, bilateral superior frontal gyrus, right postcentral gyrus, bilateral temporal fusiform cortex, left middle temporal gyrus, bilateral inferior frontal gyrus, bilateral angular gyri, right intracalcarine cortex, right supramarginal gyrus and right frontal pole, as well as in the right anterior thalamus (Fig. 1C, Supplementary Fig. 1). Furthermore, a main effect of SDB condition with medium effect size was detected in several subcortical and cortical regions including the bilateral nucleus accumbens, bilateral putamen, right central opercular cortex, bilateral frontal orbital cortex, right lateral occipital cortex, right paracingulate gyrus, and right supramarginal gyrus (Supplementary Fig. 2). Further, the SDB condition did not interact with cognitive status consistently. GMV was decreased in SDB+ relative to SDB-subjects in CU and MCI groups in general, but in AD, we observed increased GMV in the right intracalcarine cortex, right occipital fusiform gyrus, and atrophy in left middle temporal gyrus, bilateral temporal fusiform cortex, bilateral angular gyrus and the left superior frontal gyrus in SDB+ relative to SDB-subjects (Fig. 2C).

### The association between cognitive status-SDB and cognitive scores

We applied multiple linear regression models to investigate the association between regions with significant effect size of cognitive status-SDB interaction and effect sizes on MMSE scores. The variation in effect sizes on Aβ burden was linked with the variance in MMSE effect sizes (p < 0.05) in three regions: the left middle temporal gyrus, right precuneus cortex, and left occipital fusiform gyrus (Supplementary Table 1, Fig. 3A). Comparing SDB+ and SDB-conditions, we observed similar rFDG and GMV patterns in the CU and MCI groups in these three regions, but Aβ plaque burden was increased in the AD group, supported by lower CSF measures of Aβ (Fig. 3B, D).

**Fig. 3.**
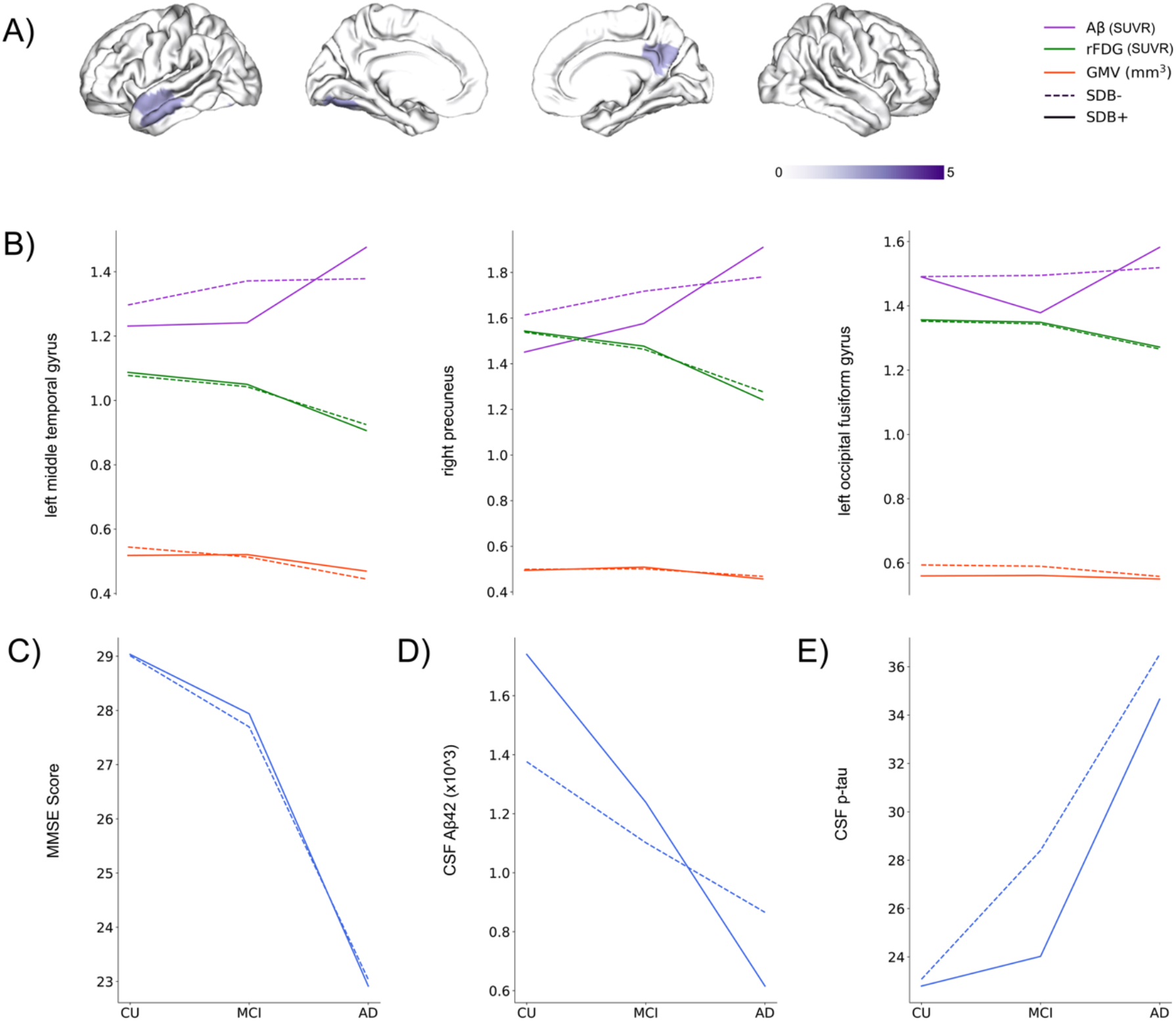
Association between cognitive status-SDB interaction and cognitive score and pattern of biomarker’s changes. A) Interaction between cognitive status (Alzheimer’s disease, mild cognitive impairment, cognitively unimpaired) and sleep-disordered breathing condition (with/without SDB) has a medium effect size on highlighted regions. Furthermore, the highlighted regions are significantly associated with cognitive status-SDB interaction effect size on mini-mental state examination (MMSE). B) brain biomarker’s changes (Aβ, rFDG, and GMV) through different cognitive status in the highlighted regions from A. C) MMSE score changes through different cognitive status. D) CSF Aβ42 changes through different cognitive status. E) CSF p-tau changes through different cognitive status.

### The association between cognitive status-SDB and CSF biomarkers

Our complementary analyses demonstrated that the effect sizes on CSF Aβ42 were associated with the effect sizes on Aβ burden in the right precuneus and PCC, with the effect sizes on rFDG in the left precuneus, and with the effect sizes on GMV in the bilateral angular gyrus and right occipital fusiform gyrus (Supplementary Fig. 3A, Supplementary Table 2). Likewise, the effect sizes on CSF p-tau were associated with the effect sizes on Aβ burden in the left lateral occipital cortex, and the effect sizes on GMV in the left middle temporal gyrus. The effect sizes on both Aβ burden and GMV in the left superior frontal gyrus were associated with the effect sizes on CSF p-tau (Supplementary Fig. 3B, Supplementary Table 3).

## Discussion

The present study assessed the moderating link between cognitive status and the presence of SDB on AD biomarkers, including Aβ, rFDG, GMV, CSF Aβ42, and CSF p-tau, across the preclinical, prodromal, and clinical AD continuum (Fig. 1, supplementary Fig. 1, 2). The direction of SDB effects was not consistent across cognitive status and brain biomarkers, with individuals with SDB showing lower Aβ burden and higher rFDG compared to those without SDB in both CU and MCI groups but higher Aβ burden and lower rFDG in the AD group (Fig. 2A, B). We found overall GMV atrophy in SDB+ individuals in the CU and MCI groups, but in the AD group, increased and decreased GMV in different regions in the SDB+ individuals were observed (Fig. 2C). The cognitive status-SDB interaction effect sizes on Aβ in the middle temporal gyrus, precuneus, and fusiform gyrus demonstrated an association with the interaction effect sizes on MMSE scores (Fig. 3A). Patterns of biomarker alterations in these three areas were pretty similar in SDB+ vs. SDB-for rFDG, GMV, and MMSE across groups (Fig. 3B, C). However, in the AD group, SDB+ subjects exhibited a higher Aβ burden in the mentioned brain areas and a steeper decline in CSF Aβ42 compared to SDB-subjects along the trajectory of disease (i.e., CU, MCI, AD), indicating the particular association of SDB-AD interaction with Aβ burden (Fig. 3B, D). The pattern for CSF p-tau also differs between SDB+ and SDB-but rather unexpectedly (less p-tau increase in SDB+). Our complementary analyses demonstrated associations between effect sizes on CSF Aβ42 and CSF p-tau with effect sizes on brain Aβ burden, rFDG, and GMV (Supplementary Fig. 3).

Collectively, these findings based on a large-scale sample demonstrate a robust association between SDB and various AD biomarkers across disease stages, with lower amyloid burden, higher functional hyperactivity, and structural atrophy being greater in preclinical and prodromal stages in SDB+ relative to SDB-individuals, while Aβ burden is particularly increased in SDB+ relative to SDB-individuals in AD. One possible explanation for these counterintuitive Aβ findings is that SDB degenerates AD-vulnerable brain networks independently of AD pathophysiology, leading to increased cognitive vulnerability to AD pathophysiology and thus resulting in earlier clinical conversion at lower overall levels of pathology. Consistent with this hypothesis, Aβ burden is lower in SDB+ individuals in preclinical and prodromal stages but higher at the end-stage diagnosis of AD. The fact that rFDG and GMV indicate hyperactivity and greater atrophy in preclinical and prodromal stages in SDB+ relative to SDB-individuals despite lower levels of Aβ burden further supports this interpretation. These effects were further associated with individual differences in CSF levels of Aβ42 and p-tau proteins and global cognitive function, with SDB status influencing the associations of Aβ burden, rFDG, and GMV with CSF biomarkers and cognitive function. Of note, the independent impact of SDB on brain atrophy has been reported previously, even in children and middle-aged adults^50–53^. SDB may thus represent a critical factor triggering greater cognitive vulnerability to AD pathophysiology.

### Interaction between cognitive status and SDB condition

The observed association between SDB and AD biomarkers is consistent with previous findings (Fig. 1A). For example, higher Aβ burden in the bilateral precuneus^54^ and temporal cortex^15^ have been reported in SDB and AD^55–57^. The previous studies demonstrated reduced rFDG uptake observed in the precuneus and superior parietal lobule in SDB^54,58^ is consistent with the medium effect size of cognitive status-SDB interaction that we observed for rFDG in AD group (Fig. 1B). Similarly, we found reduced rFDG uptake in regions such as the inferior, middle, and superior temporal gyrus, occipital gyrus, angular gyrus, precuneus, and thalamus, which are known to be AD-related regions^59^. Moreover, AD decoupled the association between regional glucose metabolism (rFDG) and functional segregation and global functional connectivity across various brain regions^60^. Our findings regarding the interaction between cognitive status and SDB in GMV (Fig. 1C) are consistent with previous reports of decreased GMV in the bilateral middle temporal gyrus in SDB^61^ and in the middle and superior temporal gyrus, parietal lobule, cingulate gyrus, thalamus, and putamen in AD^62,63^.

Considering that SDB has a differential association with Aβ (low) and rFDG (high) in CU and MCI relative to AD (high Aβ in the brain, low CSF Aβ, and low rFDG) (Fig. 2), it is possible that SDB leads to neuronal hyperactivity independent of AD pathologies, which may then contribute to cognitive impairment and decline. For example, it has been shown that poor sleep quality, shorter N2 sleep duration, and increased apnea-hypopnea index (often reported in polysomnography results of individuals with SDB) were associated with low Diffusion Tensor Imaging Analysis Along The Perivascular Space (DTI-ALPS) index of glymphatic system, lower GMV, and worse cognitive performance in community-dwelling older adults^64^. Lower DTI-ALPS is linked with higher Aβ deposition, lower CSF Aβ42, higher level of Aβ in AD^65^. Moreover, in AD, cortical accumulation of Aβ in the precuneus and PCC induces functional and structural dysconnectivity at early stages, which leads to hyperexcitability and metabolic changes and higher local tau deposition within the medial temporal lobe, resulting in widespread structural degeneration in later stages^39,66^. It has also been suggested that sleep deprivation is associated with both pathological CSF Aβ42 levels and increased Aβ plaque burden, and this relationship could be mainly associated with glymphatic system dysfunction^16^. Besides that, intracranial and intrathoracic pressure caused by apneic episodes may act as precipitating factors for neurodegeneration^16^. Overall, SDB could enhance brain vulnerability to AD pathophysiology.

### The comparison between SDB+ and SDB-groups

We found an unexpected association between the presence of SDB and lower levels of Aβ plaque burden (Fig. 2A), coupled with higher levels of rFDG uptake (Fig. 2B) in both CU and MCI individuals compared to SDB-subjects. The results from our study demonstrated that both CU and MCI individuals had higher levels of CSF Aβ42 in SDB+ compared to SDB-condition (Table 1, Fig. 3). Furthermore, previous studies suggested that lower CSF Aβ42 levels are linked to higher brain Aβ plaque burden^67,68^. Therefore, our observed low brain Aβ burden in SDB+ might be linked with high levels of CSF Aβ42 in CU and MCI groups. However, it is worth mentioning that even in individuals without SDB, 89.4% of CU, 93.5% of MCI, and 88.6% of AD group were Aβ-positive (Table 1), which may account for the similarity in Aβ levels between individuals with and without SDB in these two groups. The observed higher metabolism could indicate compensatory mechanisms or greater brain reserve^69^. However, our findings in the AD group align with previous works suggesting that SDB may modulate a complex relationship between Aβ plaque burden and rFDG metabolism^15,54,58,70^.

We observed a significant reduction of GMV in SDB+ subjects compared with SDB-individuals in various brain regions across all cognitive-status groups. Nevertheless, higher GMV for SDB+ subjects was observed in specific brain regions within all three groups. Greater GMV could reflect a higher brain reserve^69^ and be related to compensatory alterations in response to the progression of cognitive impairment to preserve cognitive performance^54^. Collectively, SDB may have a differential association with neuroimaging biomarkers in CU and MCI subjects compared to AD patients.

### The links between cognitive status-SDB interaction and cognitive scores

The similarities between CU and MCI in terms of cognitive status (measured by MMSE) might be due to the fact that our MCI subjects have similar MMSE compared to CU and were in the early stages of disease (Table 1, Fig. 3). Moreover, the pattern of MMSE score of our CU (SDB+ 29.07, SDB-29.01), MCI (SDB+ 27.93, SDB-27.69) and AD patients (SDB+ 22.94, SDB-23.04), could resemble the pattern of Aβ and rFDG changes in the mentioned three groups (Table 1, Fig. 2). The correlation between the variability of effect sizes on MMSE score and the variability of effect sizes on Aβ plaque burden in our study (Fig. 3) is consistent with findings from previous studies. For instance, visual attention, processing speed, and subtle memory impairments have been reported for subjects with SDB^58,70^. It has also been shown that SDB is associated with medial temporal lobe atrophy in older adults who are cognitively asymptomatic but on the AD continuum, which potentially heightens the risk of developing memory impairment over time^71^. Thus, we assume that by the presence of SDB, Aβ and tau contribute to synergistic neurodegenerative mechanisms, and the rate of their accumulation drastically increases^72^.

### The links between cognitive status-SDB interaction and CSF markers

Our study found a significant link between the cognitive status-SDB interaction effect sizes on CSF Aβ42 and p-tau with effect sizes of the interaction on brain Aβ (Supplementary Fig. 3A) and GMV (Supplementary Fig. 3C) in specific regions (Supplementary Table 2, Supplementary Table 3), suggesting a robust link between these CSF biomarkers and the interaction between cognitive status and SDB condition. SDB patients have shown lower CSF Aβ42 and higher total tau (t-tau)/Aβ42 ratio compared to controls and SDB patients treated by CPAP^35^. Moreover, an association has been observed between SDB and changes in CSF biomarkers of AD, specifically elevated levels of CSF Aβ42 and p-tau, in normal elderly individuals carrying the APOE e3/3 alleles^18^. These findings confirm the link between cognitive status-SDB interaction and CSF markers, which we found in this study.

### The neurobiological interaction between SDB and AD

SDB induces hypoxia, which leads to widespread maladaptive neuroinflammation by activating microglia, triggers hippocampal apoptosis, and impaired synaptic plasticity^73,74^. Intermittent hypoxia in genetically susceptible (familial) AD mice enhances Aβ plaque burden, neuroinflammation, and cognitive impairment. However, inducing chronic *sleep deprivation* impaired working memory but did not lead to neuronal dysfunction or higher Aβ accumulation in mice models^75^. This suggests that intermittent hypoxia is a key mechanism by which SDB could exacerbate AD pathology. On the other hand, higher sleep fragmentation due to SDB-induced hyperarousal prevents deep sleep and leads to slower clearance of brain wastes, particularly Aβ, through the glymphatic system (glymphatic stasis)^76,77^ (Fig. 4).

**Fig. 4.**
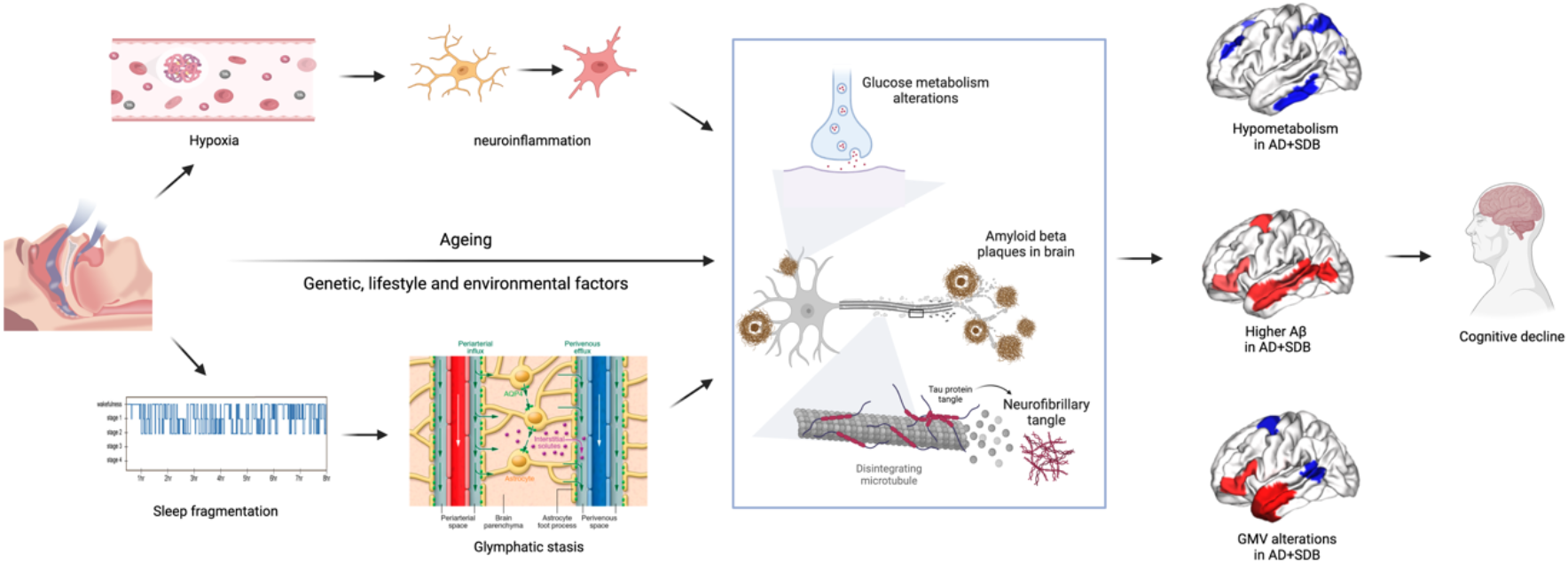
The neurobiological interaction between SDB and AD. SDB induces hypoxia and sleep fragmentation, which leads to neuroinflammation and glymphatic stasis. These changes, together with genetic, environmental, and lifestyle factors, exacerbate aging effects on the accumulation of amyloid-beta, and insoluble tau protein, which results in brain metabolic and structural alterations and precedes AD pathology.

SDB also exacerbates age-related cellular and molecular impairments, including stem cell exhaustion, telomere attrition, and epigenetic changes^78^. These changes also can alter synaptic integrity and degenerate neurons even in younger adults, who might not have Aβ in their brains yet. Put differently, these effects enhance brain vulnerability earlier than accumulation of Aβ. In later stages (i.e., AD), when subjects have higher levels of Aβ accumulation, SDB and AD have synergic maladaptive effects. These changes, together with genetic, environmental, and lifestyle factors, exacerbate aging effects on the Aβ dysregulation and increase the accumulation of amyloid-beta in the brain and raise aggregation and distribution of insoluble tau protein across the brain^74,79^. In this study, we observed that SDB is linked with higher Aβ plaque burden, reduced rFDG, and reduced/increased GMV in the AD group, but the association of SDB was prominent with both brain (in the right precuneus, left middle temporal gyrus, and left occipital fusiform gyrus) and CSF measures of Aβ plaque burden. These maladaptive processes collectively result in brain metabolic, functional, and structural alterations, which increase brain vulnerability to AD pathophysiology and lead to cognitive decline (Fig. 4).

### Limitations and future directions

Our findings should be taken with caution because of the absence of tau neuroimaging (due to lack of existing data), self-reported measures of SDB (rather than diagnosed OSA based on polysomnography), exclusion of subjects with severe SDB, who were under CPAP treatment, and finally cross-sectional design, which restrict identifying a causal relationship between SDB and AD. To address imbalanced sample size across groups, several steps were conducted to improve the stability and robustness of our results, e.g., dividing our sample into matched subsamples using stratified subsampling and running multiple iterations to calculate effect sizes. Further studies with larger and more balanced samples across different groups and longitudinal studies with multiple time points could provide more insight into dynamic changes using biomarkers to assess the interrelationship between SDB and AD over time. Further studies should investigate the effect of SDB on sleep fragmentation, tau accumulation, plasma-based biomarkers of AD, and dysfunction of the glymphatic system to reveal the underlying mechanisms of the SDB-AD interplay. The effects of SDB treatment, such as CPAP, on various neuroimaging and CSF/plasma biomarkers and cognitive outcomes in AD should also be further assessed.

## Conclusion

The present study provided evidence that SDB has a medium effect size on AD-related neuroimaging biomarkers, including Aβ, rFDG, and GMV. Individuals with SDB symptoms showed a lower Aβ plaque burden in the CU and MCI groups, but a higher Aβ plaque burden in the AD group. This could indicate that SDB increases vulnerability to the cognitive consequences of AD pathophysiology, resulting in accelerated conversion at lower levels of pathological burden in early stages. Hypermetabolism was observed in participants with SDB in the CU and MCI groups and hypometabolism in the AD group. Furthermore, SDB+ participants in the AD group exhibited altered GMV in several regions. Contrary to rFDG and GMV, the effect sizes of Aβ in the middle temporal gyrus, precuneus, and fusiform gyrus demonstrated a robust association with cognitive scores. Also, we observed that the presence of SDB results in a higher Aβ plaque burden in the above-mentioned brain regions and lower CSF Aβ42 in the AD group. Our results suggest that SDB may precipitate cognitive decline and exacerbate vulnerability to AD pathology. We hope that this multimodal study incentivizes clinicians to consider the importance of screening and treating subjects with SDB as potential therapeutic targets to reduce the burden of AD.

## Abbreviations

SDB: Sleep Disordered Breathing
AD: Alzheimer’s Disease
MCI: Mild Cognitive Disorder
CU: Cognitively Unimpaired
Aβ: Amyloid Beta plaque
rFDG: regional Fluorodeoxyglucose
GMV: Grey Matter Volume
PET: positron emission tomography
ADNI: Alzheimer’s Disease Neuroimaging Initiative
APOE: Apolipoprotein E
BMI: Body Mass Index
MMSE: Mini Mental State Examination
OSA: Obstructive Sleep Apnea

## Acknowledgments

The authors of this study would like to acknowledge the ADNI for providing the data used in this study. ADNI is a collaborative effort of many institutions and investigators, and we are grateful for their dedication and contribution to the field of Alzheimer’s disease research. We also thank the participants and their families for their willingness to participate in the ADNI project.

Data collection and sharing for this project was funded by the Alzheimer’s Disease Neuroimaging Initiative (ADNI) (National Institutes of Health Grant U01 AG024904) and DOD ADNI (Department of Defense award number W81XWH-12-2-0012). ADNI is funded by the National Institute on Aging, the National Institute of Biomedical Imaging and Bioengineering, and through generous contributions from the following: AbbVie, Alzheimer’s Association; Alzheimer’s Drug Discovery Foundation; Araclon Biotech; BioClinica, Inc.; Biogen; Bristol-Myers Squibb Company; CereSpir, Inc.; Cogstate; Eisai Inc.; Elan Pharmaceuticals, Inc.; Eli Lilly and Company; EuroImmun; F. Hoffmann-La Roche Ltd and its affiliated company Genentech, Inc.; Fujirebio; GE Healthcare; IXICO Ltd.;Janssen Alzheimer Immunotherapy Research & Development, LLC.; Johnson & Johnson Pharmaceutical Research & Development LLC.; Lumosity; Lundbeck; Merck & Co., Inc.;Meso Scale Diagnostics, LLC.; NeuroRx Research; Neurotrack Technologies; Novartis Pharmaceuticals Corporation; Pfizer Inc.; Piramal Imaging; Servier; Takeda Pharmaceutical Company; and Transition Therapeutics. The Canadian Institutes of Health Research is providing funds to support ADNI clinical sites in Canada. Private sector contributions are facilitated by the Foundation for the National Institutes of Health (www.fnih.org). The grantee organization is the Northern California Institute for Research and Education, and the study is coordinated by the Alzheimer’s Therapeutic Research Institute at the University of Southern California. ADNI data are disseminated by the Laboratory for Neuro Imaging at the University of Southern California.

