## Supplemental file for "How is sleep-disordered breathing linked with biomarkers of Alzheimer’s disease?"

### Supplementary file

#### Two-way ANCOVA detailed explanation

A two-way ANCOVA was applied on each biomarker in each region with two main factors (disease status, and SDB condition) and four covariates to control, including age, sex, APOE ε4 existence, and BMI. For each ANCOVA, we have the below factors and their degree of freedom

| | Samples | Disease status | SDB condition | Disease-SDB interaction | Age | Sex | APOE ε4 | BMI |
|---|---|---|---|---|---|---|---|---|
| Degree of freedom | 59 | 2 | 1 | 2*1=2 | 1 | 1 | 1 | 1 |

We used partial eta squared to estimate the effect size of the disease-SDB interaction on the biomarkers in different regions, as shown in Supplementary Figure 1. The null distribution differed around its mean, which was approximately 0.04. Additionally, according to the mathematics of partial eta squared, we expected the null effect size mean to be around 0.04, as shown in Equations 1[1] and 2[2].

$$\acute{\eta}_p^2 \sim \beta\left(\overbrace{\frac{df_{effect}}{2}}^{a}\ ,\ \overbrace{\frac{df_{error}}{2}}^{b}\right) \qquad , df_{\text{error}} = df_{samples} - df_{other\ factors} \qquad (1)$$

$$E(\beta) = \frac{a}{a+b} \qquad (2)$$

Similarly, when investigating only the SDB condition effect, regardless of disease status (Supplementary Figure 2), we expected the mean value for the null distribution to be around 0.02.

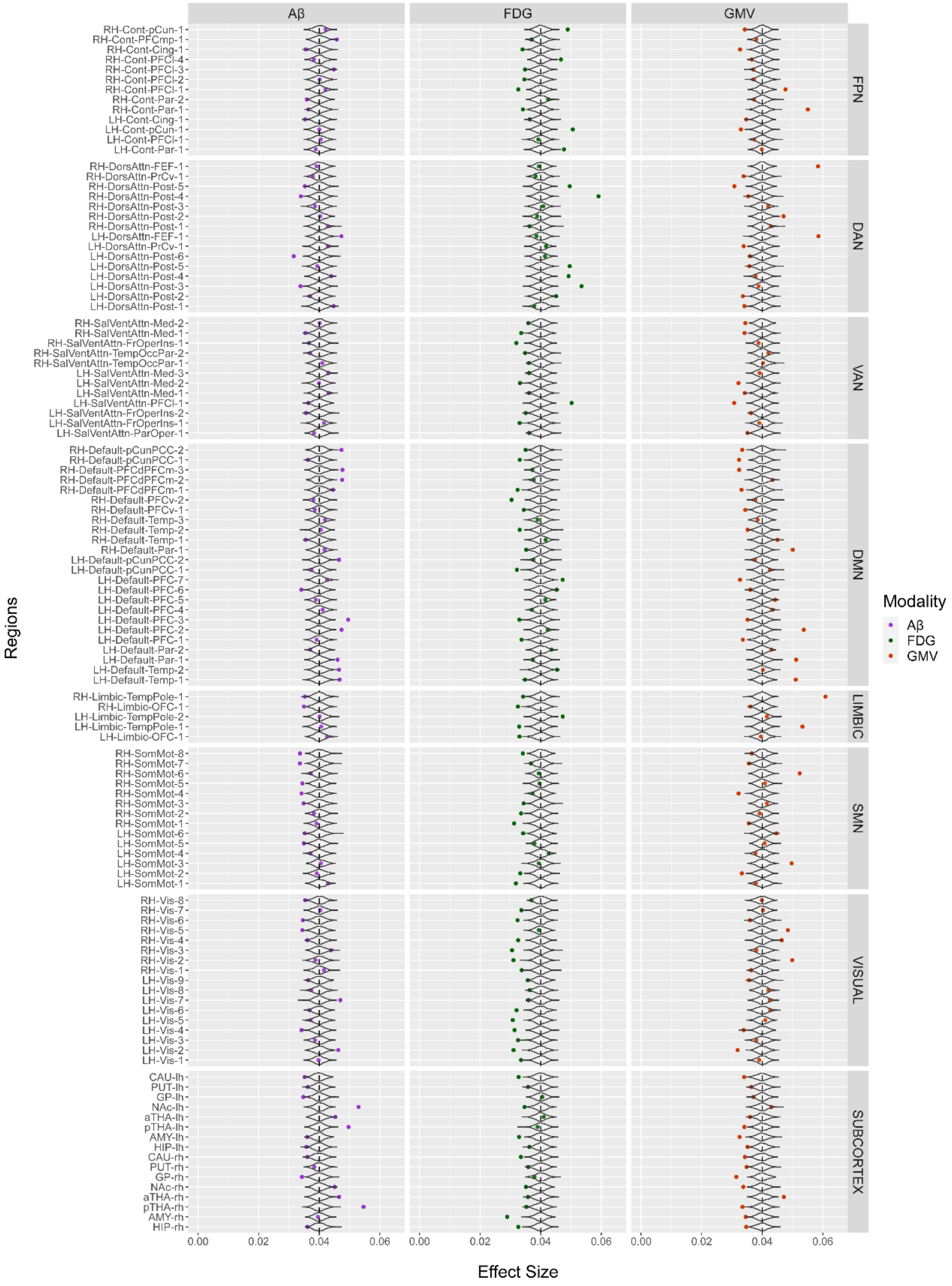


**Supplementary Figure 1. Disease-SDB interaction effect sizes on AD biomarkers.** The mean effect size of the *interaction* between disease status (Alzheimer's disease, mild cognitive impairment and cognitively unimpaired) and SDB condition (SDB-positive, and SDB-negative) for each cortical and subcortical parcel's neuroimaging biomarker (amyloid-plaque burden (Aβ), regional fluorodeoxyglucose (rFDG), grey matter volume (GMV)). The black dashed line indicates the arbitrary threshold, and null model distribution represented with a violin plot. Colored points which have passed the dashed color and are larger than the null model distribution has been considered as medium effect sizes.

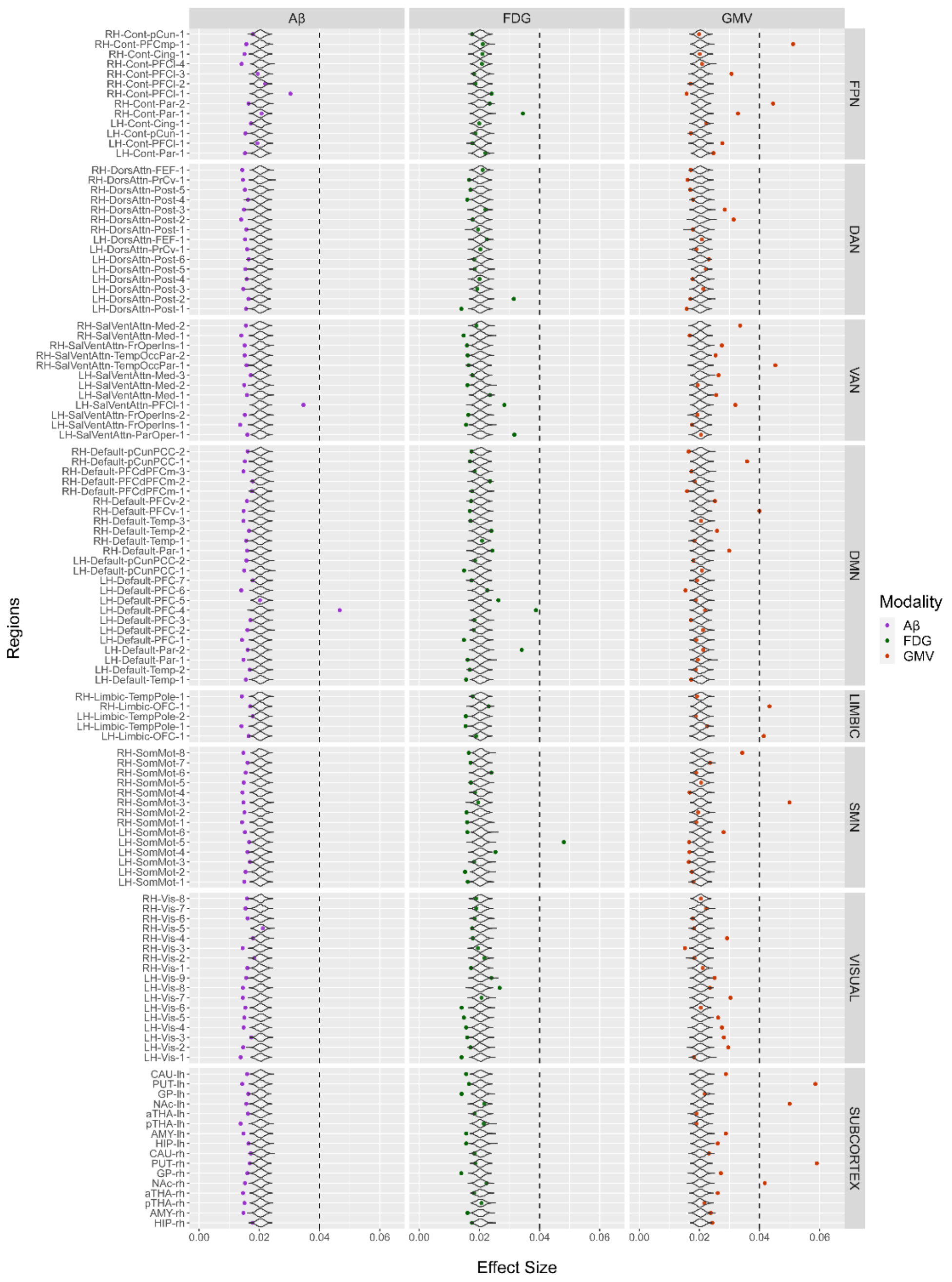


**Supplementary Figure 2. SDB condition effect sizes on AD biomarkers.** The mean effect-size of SDB condition (SDB-positive, and SDB-negative) *alone* for each cortical and subcortical parcel's neuroimaging marker (amyloid-plaque burden (Aβ), fluorodeoxyglucose (FDG), grey matter volume (GMV)). The black dashed line indicates the arbitrary threshold, and the null model distribution has represented by a violin plot. Colored points which have passed the dashed color and are larger than the null model distribution has been considered as medium effect sizes.

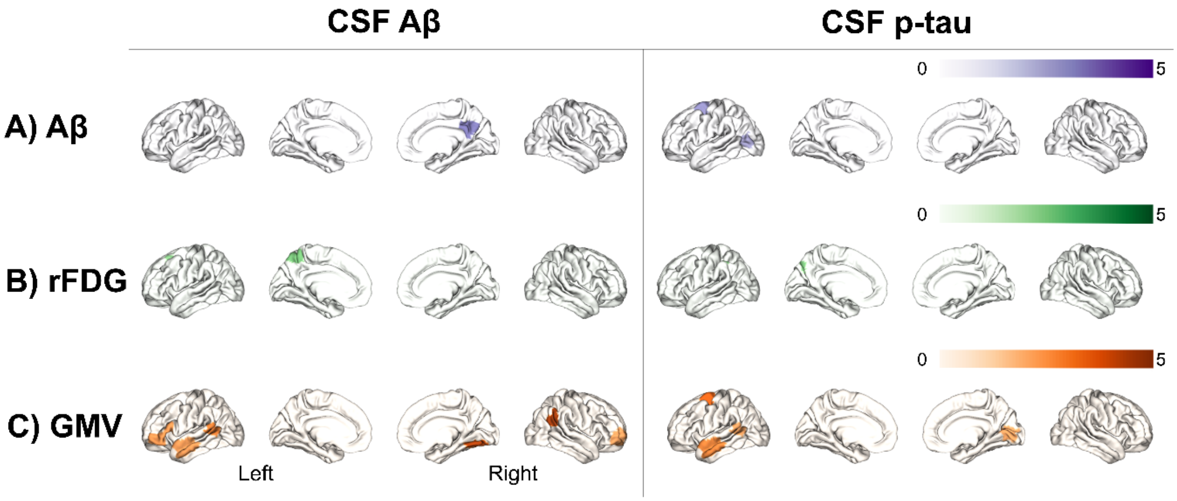


**Supplementary Figure 3. Association between disease-SDB interaction and CSF biomarkers.** Highlighted regions are significantly associated with disease-SDB interaction effect size on CSF biomarkers (CSF Aβ42, CSF p-tau).

**Supplementary Table 1.** Linear regression model findings between mini-mental state examination (MMSE) effect size of interaction between cognitive status (Alzheimer's disease, mild cognitive impairment and cognitively unimpaired) and SDB condition (with and without sleep-disordered breathing) as response variable and the effected regions, which have medium effect size (based on Fig. 1) as explanatory variables. *(*) represents significant explanatory variables.*

| Biomarker | Region | Est. | Standard Error | T-value | p-value |
|---|---|---|---|---|---|
| Aβ | Left Precuneus Cortex | 0.15 | 0.13 | 1.15 | 0.25 |
| | **Left Occipital Fusiform Gyrus** | **0.11** | **0.05** | **2.28** | **0.02*** |
| | Left Middle Temporal Gyrus | 0.1 | 0.13 | 0.8 | 0.42 |
| | Left Lateral Occipital Cortex | 0.07 | 0.09 | 0.78 | 0.43 |
| | Right Posterior Thalamus | 0.06 | 0.07 | 0.88 | 0.38 |
| | Left Nucleus Accumbens | 0.05 | 0.07 | 0.64 | 0.52 |
| | Left Cingulate Gyrus | 0.04 | 0.11 | 0.34 | 0.73 |
| | Left Inferior Frontal Gyrus | 0.03 | 0.07 | 0.43 | 0.67 |
| | Right Superior Frontal Gyrus | 0.02 | 0.08 | 0.25 | 0.8 |
| | Left Superior Frontal Gyrus | 0.01 | 0.06 | 0.08 | 0.94 |
| | Right Frontal Pole | -0.02 | 0.07 | -0.36 | 0.72 |
| | Left Posterior Thalamus | -0.05 | 0.08 | -0.66 | 0.51 |
| | Left Angular Gyrus | -0.14 | 0.12 | -1.11 | 0.27 |
| | **Left Middle Temporal Gyrus** | **-0.18** | **0.09** | **-2.09** | **0.04*** |
| | **Right Precuneus Cortex** | **-0.22** | **0.11** | **-1.98** | **0.05*** |
| rFDG | Left Precuneus Cortex | 0.04 | 0.06 | 0.68 | 0.5 |
| | Left Lateral Occipital Cortex | 0.04 | 0.07 | 0.56 | 0.57 |
| | Left Postcentral Gyrus | 0.01 | 0.05 | 0.12 | 0.9 |
| | Left Inferior Temporal Gyrus | 0.01 | 0.05 | 0.18 | 0.86 |
| | Right Lateral Occipital Cortex | 0.01 | 0.06 | 0.15 | 0.88 |
| | Right Precuneus Cortex | 0 | 0.07 | 0.03 | 0.97 |
| | Left Superior Parietal Lobule | -0.01 | 0.07 | -0.17 | 0.87 |
| | Right Middle Frontal Gyrus | -0.01 | 0.04 | -0.32 | 0.75 |
| | Left Precuneus Cortex | -0.02 | 0.07 | -0.32 | 0.75 |

| | | | | | |
|---|---|---|---|---|---|
| | Left Frontal Pole | -0.02 | 0.06 | -0.3 | 0.76 |
| | Right Superior Parietal Lobule | -0.02 | 0.06 | -0.35 | 0.73 |
| | Left Superior Frontal Gyrus | -0.04 | 0.06 | -0.57 | 0.57 |
| GMV | Left Central Opercular Cortex | 0.06 | 0.04 | 1.54 | 0.12 |
| | Right Temporal Fusiform Cortex | 0.05 | 0.04 | 1.07 | 0.28 |
| | Right Anterior Thalamus | 0.04 | 0.04 | 0.95 | 0.34 |
| | Right Intracalcarine Cortex | 0.04 | 0.04 | 1.22 | 0.22 |
| | Left Superior Frontal Gyrus | 0.03 | 0.04 | 0.75 | 0.45 |
| | Right Angular Gyrus | 0.03 | 0.04 | 0.76 | 0.45 |
| | Right Superior Frontal Gyrus | 0.02 | 0.04 | 0.4 | 0.69 |
| | Left Inferior Frontal Gyrus | 0 | 0.04 | 0.06 | 0.96 |
| | Left Angular Gyrus | -0.01 | 0.04 | -0.17 | 0.87 |
| | Right Supramarginal Gyrus | -0.01 | 0.04 | -0.24 | 0.81 |
| | Right Precentral Gyrus | -0.02 | 0.04 | -0.6 | 0.55 |
| | Right Occipital Fusiform Gyrus | -0.02 | 0.04 | -0.41 | 0.68 |
| | Right Frontal Pole | -0.04 | 0.04 | -0.91 | 0.37 |
| | Right Postcentral Gyrus | -0.04 | 0.04 | -1.01 | 0.31 |
| | Left Temporal Pole | -0.05 | 0.04 | -1.07 | 0.29 |
| | Left Middle Temporal Gyrus | -0.07 | 0.04 | -1.73 | 0.08 |

**Supplementary Table 2.** Linear regression model findings between cerebrospinal fluid amyloid-beta 42 (CSF-Aβ42) effect size of interaction between disease status (Alzheimer's disease, mild cognitive impairment and healthy control) and SDB condition (with and without sleep-disordered breathing) as response variable and the effected regions, which have medium effect size (based on Figure 2) as explanatory variables. *(*) represents significant explanatory variables.* Region labels are based on Schaefer-100 parcellation.

| Biomarker | Region | Est. | Standard Error | T-value | p-value |
|---|---|---|---|---|---|
| Aβ | LH-Vis-2 | -0.03798 | 0.083681 | -0.454 | 0.650111 |
| | LH-Vis-7 | -0.05291 | 0.156136 | -0.339 | 0.734884 |
| | LH-DorsAttn-FEF-1 | -0.16765 | 0.1081 | -1.551 | 0.121599 |
| | LH-Default-Temp-1 | -0.15419 | 0.148949 | -1.035 | 0.30112 |
| | LH-Default-Temp-2 | 0.33224 | 0.21691 | 1.532 | 0.126275 |
| | LH-Default-Par-1 | 0.051915 | 0.205687 | 0.252 | 0.800845 |
| | LH-Default-PFC-2 | -0.10194 | 0.11196 | -0.91 | 0.363049 |
| | LH-Default-PFC-3 | -0.01948 | 0.179244 | -0.109 | 0.913489 |
| | LH-Default-pCunPCC-2 | -0.04921 | 0.224227 | -0.219 | 0.826396 |
| | RH-Default-PFCdPFCm-2 | -0.05388 | 0.112498 | -0.479 | 0.632234 |
| | RH-Default-PFCdPFCm-3 | -0.02148 | 0.133863 | -0.16 | 0.872617 |
| | **RH-Default-pCunPCC-2** | **0.497442** | **0.193063** | **2.577** | **0.010284*** |
| | pTHA-lh | 0.091159 | 0.136083 | 0.67 | 0.503267 |
| | NAc-lh | -0.12881 | 0.121517 | -1.06 | 0.28967 |
| | pTHA-rh | -0.14733 | 0.142632 | -1.033 | 0.30218 |
| FDG | LH-DorsAttn-Post-3 | 0.148146 | 0.120521 | 1.229 | 0.219612 |
| | LH-DorsAttn-Post-4 | 0.05369 | 0.078963 | 0.68 | 0.496879 |
| | **LH-DorsAttn-Post-5** | **-0.22373** | **0.112403** | **-1.99** | **0.047122*** |
| | LH-SalVentAttn-PFCl-1 | -0.13149 | 0.103054 | -1.276 | 0.202621 |
| | LH-Limbic-TempPole-2 | 0.072824 | 0.081422 | 0.894 | 0.371569 |
| | LH-Cont-Par-1 | -0.1099 | 0.120543 | -0.912 | 0.36239 |
| | LH-Cont-pCun-1 | 0.169778 | 0.105097 | 1.615 | 0.106891 |

| | | | | | |
|---|---|---|---|---|---|
| | LH-Default-PFC-7 | 0.187169 | 0.105842 | 1.768 | 0.077652 |
| | RH-DorsAttn-Post-4 | 0.005778 | 0.097272 | 0.059 | 0.952659 |
| | RH-DorsAttn-Post-5 | 0.045968 | 0.102115 | 0.45 | 0.652801 |
| | RH-Cont-PFCl-4 | 0.036623 | 0.074259 | 0.493 | 0.622114 |
| | RH-Cont-pCun-1 | -0.05298 | 0.118016 | -0.449 | 0.653695 |
| GMV | LH-SomMot-3 | -0.02794 | 0.060799 | -0.459 | 0.646094 |
| | LH-DorsAttn-FEF-1 | -0.08079 | 0.062484 | -1.293 | 0.196649 |
| | LH-Limbic-TempPole-1 | -0.07086 | 0.074177 | -0.955 | 0.339922 |
| | LH-Default-Temp-1 | -0.11685 | 0.065949 | -1.772 | 0.077085 |
| | **LH-Default-Par-1** | **0.13821** | **0.0637** | **2.17** | **0.030534*** |
| | LH-Default-PFC-2 | 0.111411 | 0.06092 | 1.829 | 0.068066 |
| | **RH-Vis-2** | **-0.22414** | **0.068245** | **-3.284** | **0.001099*** |
| | RH-Vis-5 | 0.021723 | 0.059128 | 0.367 | 0.713492 |
| | RH-SomMot-6 | 0.074479 | 0.068594 | 1.086 | 0.278135 |
| | RH-DorsAttn-Post-2 | -0.01901 | 0.065891 | -0.288 | 0.773107 |
| | RH-DorsAttn-FEF-1 | 0.007002 | 0.067522 | 0.104 | 0.917456 |
| | RH-Limbic-TempPole-1 | 0.070916 | 0.071552 | 0.991 | 0.322147 |
| | RH-Cont-Par-1 | -0.09106 | 0.068424 | -1.331 | 0.183881 |
| | RH-Cont-PFCl-1 | -0.11583 | 0.069764 | -1.66 | 0.097528 |
| | **RH-Default-Par-1** | **0.224837** | **0.064812** | **3.469** | **0.000571*** |
| | aTHA-rh | 0.054284 | 0.064756 | 0.838 | 0.402301 |

**Supplementary Table 3.** Linear regression model findings between cerebrospinal fluid phosphorylated tau (CSF-ptau) effect size of interaction between disease status (Alzheimer's disease, mild cognitive impairment and healthy control) and SDB condition (with and without sleep-disordered breathing) as response variable and the effected regions, which have medium effect size (based on Figure 2) as explanatory variables. *(*) represents significant explanatory variables.* Region labels are based on Schaefer-100 parcellation.

| Biomarker | Region | Est. | Standard Error | T-value | p-value |
|---|---|---|---|---|---|
| Aβ | LH-Vis-2 | -0.06027 | 0.054585 | -1.104 | 0.27011 |
| | **LH-Vis-7** | **0.20152** | **0.101849** | **1.979** | **0.04845** |
| | **LH-DorsAttn-FEF-1** | **-0.15907** | **0.070514** | **-2.256** | **0.02454** |
| | LH-Default-Temp-1 | -0.01814 | 0.09716 | -0.187 | 0.85196 |
| | LH-Default-Temp-2 | 0.118438 | 0.141492 | 0.837 | 0.40298 |
| | LH-Default-Par-1 | -0.15522 | 0.134171 | -1.157 | 0.24791 |
| | LH-Default-PFC-2 | 0.054352 | 0.073032 | 0.744 | 0.45712 |
| | LH-Default-PFC-3 | 0.033723 | 0.116922 | 0.288 | 0.77315 |
| | LH-Default-pCunPCC-2 | 0.131573 | 0.146265 | 0.9 | 0.36882 |
| | RH-Default-PFCdPFCm-2 | -0.01234 | 0.073383 | -0.168 | 0.86651 |
| | RH-Default-PFCdPFCm-3 | 0.093567 | 0.08732 | 1.072 | 0.28448 |
| | RH-Default-pCunPCC-2 | -0.15998 | 0.125936 | -1.27 | 0.2046 |
| | pTHA-lh | -0.00255 | 0.088768 | -0.029 | 0.97706 |
| | NAc-lh | -0.00782 | 0.079266 | -0.099 | 0.9215 |
| | pTHA-rh | 0.033481 | 0.09304 | 0.36 | 0.71911 |
| FDG | LH-DorsAttn-Post-3 | 0.088771 | 0.078617 | 1.129 | 0.25941 |
| | LH-DorsAttn-Post-4 | -0.00671 | 0.051508 | -0.13 | 0.89635 |
| | LH-DorsAttn-Post-5 | -0.1058 | 0.073321 | -1.443 | 0.14969 |
| | LH-SalVentAttn-PFCl-1 | -0.03356 | 0.067223 | -0.499 | 0.61789 |
| | LH-Limbic-TempPole-2 | -0.01298 | 0.053112 | -0.244 | 0.80709 |
| | **LH-Cont-Par-1** | **-0.15012** | **0.078631** | **-1.909** | **0.05685** |
| | **LH-Cont-pCun-1** | **0.132468** | **0.068556** | **1.932** | **0.05393** |

| | | | | | |
|---|---|---|---|---|---|
| | LH-Default-PFC-7 | 0.034715 | 0.069042 | 0.503 | 0.61534 |
| | RH-DorsAttn-Post-4 | -0.04486 | 0.063451 | -0.707 | 0.47989 |
| | RH-DorsAttn-Post-5 | 0.099149 | 0.06661 | 1.488 | 0.1373 |
| | RH-Cont-PFCl-4 | -0.01039 | 0.048439 | -0.214 | 0.83025 |
| | RH-Cont-pCun-1 | -0.06995 | 0.076983 | -0.909 | 0.36402 |
| GMV | LH-SomMot-3 | -0.03153 | 0.039659 | -0.795 | 0.42703 |
| | **LH-DorsAttn-FEF-1** | **-0.1084** | **0.040759** | **-2.66** | **0.00809** |
| | LH-Limbic-TempPole-1 | 0.008093 | 0.048386 | 0.167 | 0.86724 |
| | **LH-Default-Temp-1** | **0.089886** | **0.043019** | **2.089** | **0.03721** |
| | LH-Default-Par-1 | 0.068593 | 0.041552 | 1.651 | 0.09946 |
| | LH-Default-PFC-2 | 0.005357 | 0.039738 | 0.135 | 0.89282 |
| | RH-Vis-2 | 0.026761 | 0.044517 | 0.601 | 0.54803 |
| | RH-Vis-5 | 0.063798 | 0.03857 | 1.654 | 0.09878 |
| | RH-SomMot-6 | -0.02953 | 0.044745 | -0.66 | 0.50955 |
| | RH-DorsAttn-Post-2 | 0.021359 | 0.042981 | 0.497 | 0.61947 |
| | RH-DorsAttn-FEF-1 | 0.043181 | 0.044045 | 0.98 | 0.32741 |
| | RH-Limbic-TempPole-1 | -0.01941 | 0.046674 | -0.416 | 0.67765 |
| | RH-Cont-Par-1 | 0.008203 | 0.044634 | 0.184 | 0.85427 |
| | RH-Cont-PFCl-1 | -0.05378 | 0.045508 | -1.182 | 0.23787 |
| | RH-Default-Par-1 | 0.040352 | 0.042277 | 0.954 | 0.34034 |
| | aTHA-rh | 0.046348 | 0.042241 | 1.097 | 0.2731 |
